# Parkinson’s Progression Markers Initiative: A Milestone-Based Strategy to Monitor PD Progression

**DOI:** 10.1101/2023.05.23.23290344

**Authors:** Michael C. Brumm, Andrew Siderowf, Tanya Simuni, Elliot Burghardt, Seung Ho Choi, Chelsea Caspell-Garcia, Lana M. Chahine, Brit Mollenhauer, Tatiana Foroud, Douglas Galasko, Kalpana Merchant, Vanessa Arnedo, Samantha J. Hutten, Alyssa N. O’Grady, Kathleen L. Poston, Caroline M. Tanner, Daniel Weintraub, Karl Kieburtz, Kenneth Marek, Christopher S. Coffey Parkinson’s Progression Markers Initiative

## Abstract

**Background:** Identifying a meaningful progression metric for Parkinson’s disease (PD) that reflects heterogeneity remains a challenge.

**Objective:** To assess the frequency and baseline predictors of progression to clinically relevant motor and non-motor PD milestones.

**Methods:** Using data from the Parkinson’s Progression Markers Initiative (PPMI) *de novo* PD cohort, we monitored 25 milestones across six domains (“walking and balance”; “motor complications”; “cognition”; “autonomic dysfunction”; “functional dependence”; “activities of daily living”). Milestones were intended to be severe enough to reflect meaningful disability. We assessed the proportion of participants reaching any milestone; evaluated which occurred most frequently; and conducted a time-to-first-event analysis exploring whether baseline characteristics were associated with progression.

**Results:** Half of participants reached at least one milestone within five years. Milestones within the cognitive, functional dependence, and autonomic dysfunction domains were reached most often. Among participants who reached a milestone at an annual follow-up visit and remained active in the study, 82% continued to meet criteria for any milestone at one or more subsequent annual visits and 55% did so at the *next* annual visit. In multivariable analysis, baseline features predicting faster time to reaching a milestone included age (*p*<0.0001), greater MDS-UPDRS total scores (*p*<0.0001), higher GDS-15 depression scores (*p*=0.0341), lower dopamine transporter binding (*p*=0.0043), and lower CSF total α-synuclein levels (*p*=0.0033). Symptomatic treatment was not significantly associated with reaching a milestone (*p*=0.1639).

**Conclusions:** Clinically relevant milestones occur frequently, even in early PD. Milestones were significantly associated with baseline clinical and biological markers, but not with symptomatic treatment. Further studies are necessary to validate these results, further assess the stability of milestones, and explore translating them into an outcome measure suitable for observational and therapeutic studies.

## INTRODUCTION

The progressive course and diverse motor and non-motor features of Parkinson’s disease (PD) have been recognized since the earliest descriptions of the disorder [1]. Although PD is classically defined based on cardinal motor features, cognitive decline and a spectrum of other non-motor features may emerge and progress along the disease course and result in substantial disability [2-6]. Identifying a clinically meaningful progression metric for testing novel therapeutics that reflects this heterogeneity has proven to be a challenge. Several different ways of defining progression have been implemented as outcomes in trials based on motor, cognitive, or biomarker outcomes [1, 7, 8]. However, none have been entirely satisfactory for either confirming or rejecting putative disease-modifying effects because they fail to capture the protean features that progressive PD can produce. Defining progression has also proven difficult for observational and biomarker verification studies utilizing Parkinson’s Progression Markers Initiative (PPMI) data and specimens, with challenges including differences in ON vs. OFF state data completeness patterns among sporadic vs. genetic PD cohorts [9] and evidence that PD participants who dropped out early had lower cognitive performance at their last completed visit [10]. Thus, a challenge for future PD research is to develop reliable and valid endpoints that can account for progression across the spectrum of clinical features and are versatile in the context of incomplete data.

Change in the Unified Parkinson’s Disease Rating Scale (UPDRS) [11] and Movement Disorder Society UPDRS (MDS-UPDRS) [12] have been the most common metrics for quantifying disease progression [13, 14]. While the MDS-UPDRS has been useful for testing symptomatic drugs, several limitations have been recognized. First, only Part II measures functional outcomes and is thus intrinsically clinically meaningful. Second, the MDS-UPDRS, especially the motor examination (Part III), is highly sensitive to the impact of symptomatic treatment [15]. As a result, disease-modifying therapies have typically been tested during the brief period between diagnosis and the initiation of symptomatic treatment and only progression of motor disability may be assessed. In this paradigm, only a small fraction of PD patients is eligible to participate in disease-modifying trials, and participants must often trade-off between the need for symptomatic treatment and trial participation.

An alternative approach is to record the emergence of clinically relevant outcomes. This approach is accepted in other fields of medicine. For example, in therapies for vascular disease, composite outcomes combining mortality with nonfatal events—including myocardial infarction, stroke, and revascularization—are widely used and considered to be a measure of clinically meaningful impacts of the disease [16]. We sought to identify a similar approach to measuring progression in PD patients as they move from diagnosis into the middle stages of disease when disability becomes more apparent. We utilized PPMI data to define and measure a composite endpoint comprised of 25 “progression milestones” spanning six domains. These components were selected based on expert consensus to reflect meaningful PD disability such that meeting a milestone would represent unequivocal disease progression. Primary analyses assessed the frequency of reaching any milestone within a five-year follow-up period after enrollment and explored whether baseline factors—including demographic characteristics, clinical features of PD, and cerebrospinal fluid (CSF) and imaging biomarkers—were associated with time to progression. In addition, sample size estimates were calculated to evaluate proof-of-concept and provide a benchmark for future efforts to refine this framework for possible use in therapeutic trials.

## METHODS

### Study sample

PPMI is a multicenter, international, prospective cohort study. Study aims and methodology have been published elsewhere [17]. Study protocol and manuals are available at https://www.ppmi-info.org/study-design. PPMI sites received approval from an ethical standards committee on human experimentation before study initiation and obtained written informed consent for research from all participants in the study. PD participants included in this analysis were recently diagnosed (mean [SD] duration from diagnosis: 6.6 [6.5] months) and untreated with PD medications at the time of enrollment. Participants were required to be aged 30 years or older (at diagnosis); have a Hoehn and Yahr score of < 3; and either have two symptoms out of resting tremor, bradykinesia, or rigidity (including either resting tremor or bradykinesia), or asymmetric resting tremor or asymmetric bradykinesia. In addition, all participants underwent a screening dopamine transporter (DAT) or vesicular monoamine transporter (VMAT) scan and were required to have evidence of dopaminergic deficit consistent with PD.

### Assessments

#### Baseline Measures

All participants underwent a comprehensive baseline evaluation—including clinical testing, imaging assessments, and biospecimen collection—as detailed elsewhere [18]. From these data, a pre-specified set of candidate predictor variables were considered for this analysis. This encompassed demographics, including age, sex, and clinical site (US vs. non-US); body mass index (kg/m^2^); orthostatic (supine to standing) change in systolic blood pressure; and duration of disease (months from diagnosis). Clinical assessments of motor and non-motor PD characteristics comprised the MDS-UPDRS, including Hoehn and Yahr stage and derived tremor and postural instability/gait difficulty (PIGD) scores [12, 19]; modified Schwab and England Activities of Daily Living Scale (S&E); Montreal Cognitive Assessment (MoCA); Scales for Outcomes in Parkinson’s Disease-Autonomic (SCOPA-AUT); 15-item Geriatric Depression Scale (GDS-15); State-Trait Anxiety Inventory (STAI); Epworth Sleepiness Scale (ESS); REM Sleep Behavior Disorder (RBD) Screening Questionnaire (RBDSQ); and University of Pennsylvania Smell Identification Test (UPSIT). Lastly, selected biomarker variables included two dopamine transporter (DAT) specific binding ratio (SBR) measures, mean striatum SBR and mean putamen SBR; serum uric acid (urate); and four CSF biomarkers: CSF total α-synuclein (α-syn) was measured using a sandwich-type immunoassay kit (BioLegend; formerly Covance) [20] and CSF amyloid beta (Aβ_1−42_), total tau (*t*-tau), and phosphorylated tau_181_ (*p*-tau) were measured using Elecsys electrochemiluminescence immunoassays (Roche Diagnostics) [21].

#### Longitudinal Measures

Time to initiation of PD medication was determined based on the initiation date of symptomatic treatment for motor features of PD, as previously described [22]. Standard clinical metrics were assessed at least annually after baseline (see Table 1). This included the MDS-UPDRS, Hoehn & Yahr stage, MoCA, SCOPA-AUT, S&E, and blood pressure (supine and standing) measurements. Additional measures included the standardized results (per published norms) of a detailed cognitive battery, described previously [23], evaluating four cognitive domains: memory (assessed by the Hopkins Verbal Learning Test-Revised [HVLT-R] immediate free recall [i.e., total recall] and recognition discrimination index scores); visuospatial function (Benton Judgment of Line Orientation 15-item [split-half] version); processing speed-attention (Symbol-Digit Modalities Test); and executive function and working memory (Letter-Number Sequencing and semantic [animal] fluency). Furthermore, cognitive categorization assessments completed annually by PPMI site investigators yielded two variables of interest: (1) clinical diagnosis of PD dementia (PDD) [24]; and (2) presence of significant functional impairment due to cognitive deficits.

**Table 1.**
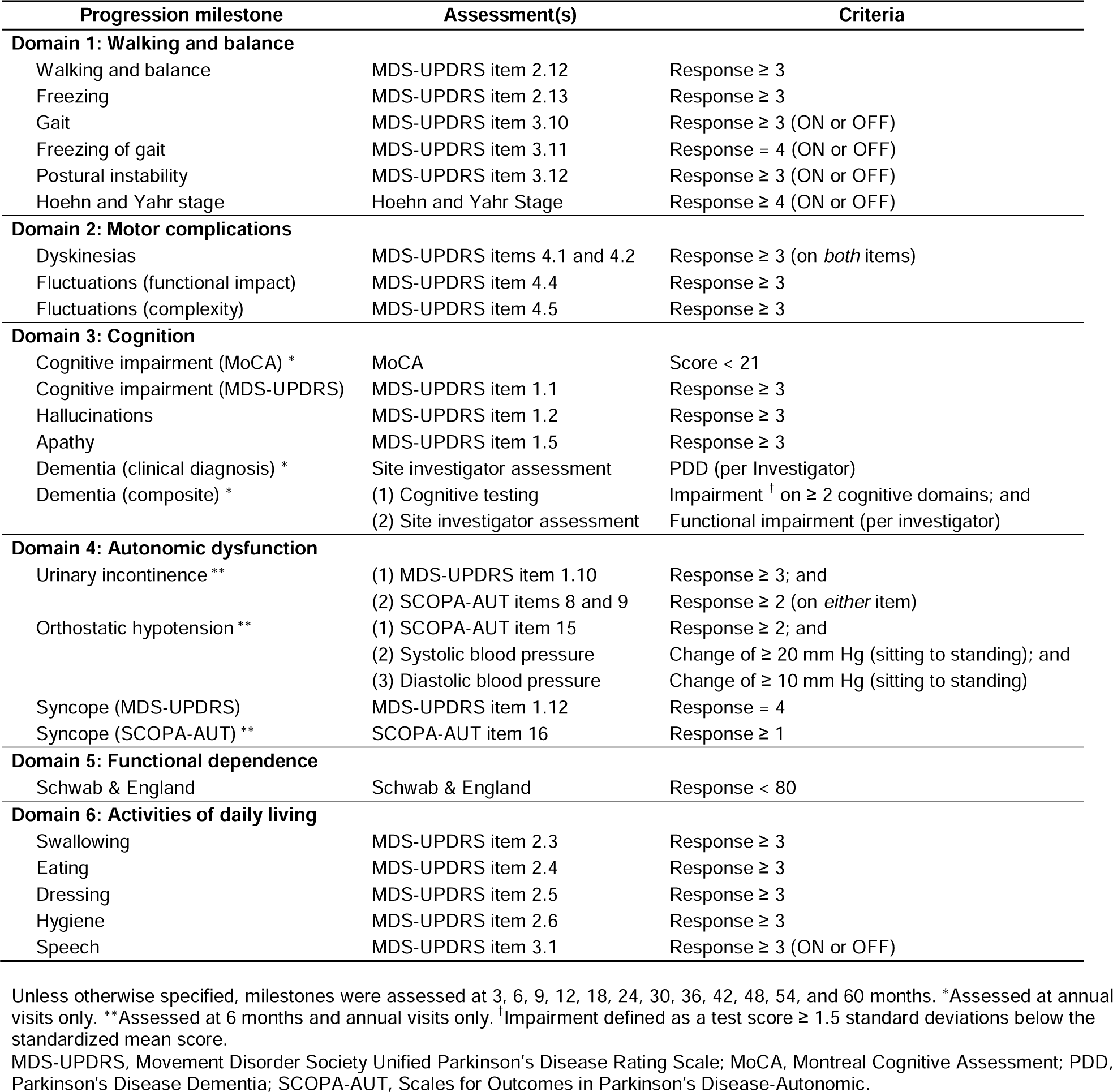
Criteria used to define progression milestones

| Progression milestone | Assessment(s) | Criteria |
| --- | --- | --- |
| **Domain 1: Walking and balance** |  |  |
| Walking and balance | MDS-UPDRS item 2.12 | Response $\geq 3$ |
| Freezing | MDS-UPDRS item 2.13 | Response $\geq 3$ |
| Gait | MDS-UPDRS item 3.10 | Response $\geq 3$ (ON or OFF) |
| Freezing of gait | MDS-UPDRS item 3.11 | Response = 4 (ON or OFF) |
| Postural instability | MDS-UPDRS item 3.12 | Response $\geq 3$ (ON or OFF) |
| Hoehn and Yahr stage | Hoehn and Yahr Stage | Response $\geq 4$ (ON or OFF) |
| **Domain 2: Motor complications** |  |  |
| Dyskinesias | MDS-UPDRS items 4.1 and 4.2 | Response $\geq 3$ (on *both* items) |
| Fluctuations (functional impact) | MDS-UPDRS item 4.4 | Response $\geq 3$ |
| Fluctuations (complexity) | MDS-UPDRS item 4.5 | Response $\geq 3$ |
| **Domain 3: Cognition** |  |  |
| Cognitive impairment (MoCA) * | MoCA | Score < 21 |
| Cognitive impairment (MDS-UPDRS) | MDS-UPDRS item 1.1 | Response $\geq 3$ |
| Hallucinations | MDS-UPDRS item 1.2 | Response $\geq 3$ |
| Apathy | MDS-UPDRS item 1.5 | Response $\geq 3$ |
| Dementia (clinical diagnosis) * | Site investigator assessment | PDD (per Investigator) |
| Dementia (composite) * | (1) Cognitive testing<br>(2) Site investigator assessment | Impairment <sup>†</sup> on $\geq 2$ cognitive domains; and<br>Functional impairment (per investigator) |
| **Domain 4: Autonomic dysfunction** |  |  |
| Urinary incontinence ** | (1) MDS-UPDRS item 1.10<br>(2) SCOPA-AUT items 8 and 9 | Response $\geq 3$; and<br>Response $\geq 2$ (on *either* item) |
| Orthostatic hypotension ** | (1) SCOPA-AUT item 15<br>(2) Systolic blood pressure<br>(3) Diastolic blood pressure | Response $\geq 2$; and<br>Change of $\geq 20$ mm Hg (sitting to standing); and<br>Change of $\geq 10$ mm Hg (sitting to standing) |
| Syncope (MDS-UPDRS) | MDS-UPDRS item 1.12 | Response = 4 |
| Syncope (SCOPA-AUT) ** | SCOPA-AUT item 16 | Response $\geq 1$ |
| **Domain 5: Functional dependence** |  |  |
| Schwab & England | Schwab & England | Response < 80 |
| **Domain 6: Activities of daily living** |  |  |
| Swallowing | MDS-UPDRS item 2.3 | Response $\geq 3$ |
| Eating | MDS-UPDRS item 2.4 | Response $\geq 3$ |
| Dressing | MDS-UPDRS item 2.5 | Response $\geq 3$ |
| Hygiene | MDS-UPDRS item 2.6 | Response $\geq 3$ |
| Speech | MDS-UPDRS item 3.1 | Response $\geq 3$ (ON or OFF) |

Unless otherwise specified, milestones were assessed at 3, 6, 9, 12, 18, 24, 30, 36, 42, 48, 54, and 60 months. \*Assessed at annual visits only. \*\*Assessed at 6 months and annual visits only. <sup>†</sup>Impairment defined as a test score $\geq 1.5$ standard deviations below the standardized mean score.
MDS-UPDRS, Movement Disorder Society Unified Parkinson's Disease Rating Scale; MoCA, Montreal Cognitive Assessment; PDD, Parkinson's Disease Dementia; SCOPA-AUT, Scales for Outcomes in Parkinson's Disease-Autonomic.

#### Progression Milestones

From the longitudinal assessments described above, criteria were established for 25 progression milestones spanning six clinical domains: “walking and balance”; “motor complications”; “cognition”; “autonomic dysfunction”; “functional dependence”; and “activities of daily living.” Table 1 lists all milestones, grouped by domain, and specifies the criteria by which they were defined. Milestones were chosen by a working group of clinical experts (AS, TS, LMC, BM, DG, KLP, CMT, DW, KK, KM) based on knowledge of the existing literature [2-6, 25] and clinical experience. This process included several sequential steps. First, the working group convened for a series of meetings and agreed on an overarching strategy of defining progression using a multidimensional composite endpoint. Second, the same panel reviewed the rating scales and other outcome assessments included in the PPMI protocol and identified items that measured dysfunction within the dimensions of interest (e.g., motor, cognitive, autonomic); in doing so, a concerted effort was made to omit items that are particularly sensitive to the effects of symptomatic therapy (e.g., MDS-UPDRS Part III items measuring tremor). Third, in cases where scale items had multiple levels, the panel agreed upon levels that represented unequivocal and at least moderately severe forms of the type of disability they were intended to capture and reflected a degree of dysfunction that is recognized as clinically meaningful within the expert community. Lastly, to facilitate interpretability by grouping milestones into categories that were consistent with clinical practice, components of the composite endpoint were classified across six clinical domains.

Per protocol, most milestones were assessed quarterly for one year and semiannually thereafter; however, three autonomic dysfunction milestones were only assessed at six months and then annually, and three cognitive milestones were only assessed annually. As previously described [23, 26], the site investigator’s determination of cognitive impairment (from which two dementia-related milestones were derived) was introduced after some participants had already completed their baseline and 12-month visits; consequently, most PD participants (74.9%) missed this assessment at baseline and roughly a third missed it at 12 months. Otherwise, missing data were rare. In all instances of missing data, a conservative approach was applied by which it was assumed that the corresponding milestone criteria were *not* met.

A composite binary endpoint, defined as time to first occurrence of any one of the milestones, comprised the primary outcome variable. Participants who met milestone criteria at baseline and/or never completed any follow-up visits were excluded.

### Data Sources

Using archived data (downloaded from https://www.ppmi-info.org/access-data-specimens/download-data and reflecting data captured in the PPMI database as of June 30, 2020; RRID:SCR_006431), two analysis data sets were derived. The first data set computed the primary endpoint based on data collected at the first five *annual* follow-up visits only (i.e., the visits at which *all* milestones were evaluated per protocol). A second data set derived the primary endpoint from the first five annual follow-up visits *and* seven additional “interim” visits (scheduled at 3, 6, 9, 18, 30, 42, and 54 months). Interim visits evaluated most, but not all, progression milestones. To gauge the possible implications of the frequency of endpoint assessments on future study design, most analyses evaluated both data sets. However, for ease of interpretation and to ensure equal weighting across milestones, models examining baseline predictors of time-to-progression were fitted using *annual* data only.

### Statistical Analysis

Figures were created using RStudio (Posit Software, PBC, Boston, MA; <u>posit.co</u>; RRID:SCR_000432) [27]. All other analyses were performed using SAS v9.4 (SAS Institute Inc., Cary, NC; <u>sas.com</u>; RRID:SCR_008567). To identify baseline predictors of progression, a time-to-event analysis was conducted using multivariable Cox proportional hazard models with a backward selection approach. Time was calculated from the date of enrollment until the date of the first annual visit at which criteria for at least one milestone were met. Participants who never met milestone criteria were censored at the time of their last completed annual visit. Participants who met criteria for any milestone at baseline and/or did not complete at least one annual follow-up were excluded from all models. Ties were handled using Efron’s approximation. For model fitting, a covariate was included if it was associated with time to progression at a significance level of 0.10 or less. PD medication use (i.e., a binary indicator variable for whether symptomatic therapy had been initiated) was included as a time-dependent covariate. Due to skewed distributions, rank values were used for all CSF biomarker variables. For CSF total α-syn, all values were included regardless of hemoglobin level; however, sensitivity analyses were conducted that excluded samples with hemoglobin levels exceeding 200 ng/mL [28]. To address multicollinearity during model selection, the MDS-UPDRS total score was considered for the multivariable model instead of Hoehn & Yahr stage and PIGD Score. Similarly, among biomarker variables, mean striatum SBR was prioritized over mean putamen SBR and the ratio of CSF *t*-tau/Aβ_1−42_ was favored over CSF Aβ_1−42_ alone. This screening process revealed a set of potential predictor variables, which made up an initial “full model.” Subsequently, a backward selection process removed variables one at a time until all variables remaining in the model were significant at the 0.05 level. For all steps in the backward selection process, sex and PD medication use were forced into the model. Due to the exploratory nature of these analyses, no adjustments were made for multiple comparisons.

As secondary analyses, we performed sample size calculations for a hypothetical trial targeting 80% power for a two-sided log-rank test (α = 0.05) comparing the survival curves of two treatment groups using a balanced design. Variable assumptions included study length (two vs. three years) and the hazard ratio of the experimental group relative to the comparison group (0.50 vs. 0.75). The comparison group’s survival curve was approximated using a piecewise linear curve based on survival function estimates derived from two separate data sources (the “annual visits” vs. “all visits” data sets defined above). Survival function estimates were computed using Kaplan-Meier estimators, with time rounded to the nearest 3 months (i.e., per-protocol time).

## RESULTS

Supplementary Figure 1 presents a flow chart summarizing how many participants were assessed at each study time point. Out of 423 PD participants enrolled, 32 (7.6%) met criteria at baseline for at least one progression milestone. This included two participants who met baseline criteria within two domains (in one case, autonomic dysfunction and walking and balance; in the other, autonomic dysfunction and activities of daily living) and 30 who did so for one domain only (13 autonomic dysfunction, five walking and balance, five cognition, four activities of daily living, three functional dependence). These participants were excluded from all analyses. The remaining 391 participants had a median duration of follow-up of seven years and a 5-year dropout rate of 18%. Finally, 6/391 participants never completed any follow-up visits and were excluded from additional analyses. Supplementary Table 1 presents baseline demographic and disease characteristics for the remaining participants.

Figure 1 depicts Kaplan-Meier estimates of progression-free survival. The first curve, derived from *annual* milestone assessments only (at 12, 24, 36, 48, and 60 months), reflects 376 participants who were milestone-free at baseline and subsequently completed at least one annual follow-up. Of these participants, 166 (44.1%) met criteria for the composite endpoint at one or more annual visits. Kaplan-Meier estimates of the 12, 24, and 36-month progression rates equaled 10.1%, 20.8%, and 30.6%, respectively. By contrast, the second curve is derived from milestone data collected across the first five annual visits *and* seven additional interim visits (3, 6, 9, 18, 30, 42, and 54 months). Out of 385 participants who were milestone-free at baseline and returned for at least one follow-up assessment (annual or interim), 193 (50.1%) met progression milestone criteria during at least one of these 12 assessments, with corresponding 12, 24, and 36-month progression rates of 16.6%, 27.7%, and 37.4%, respectively.

**Fig. 1.**
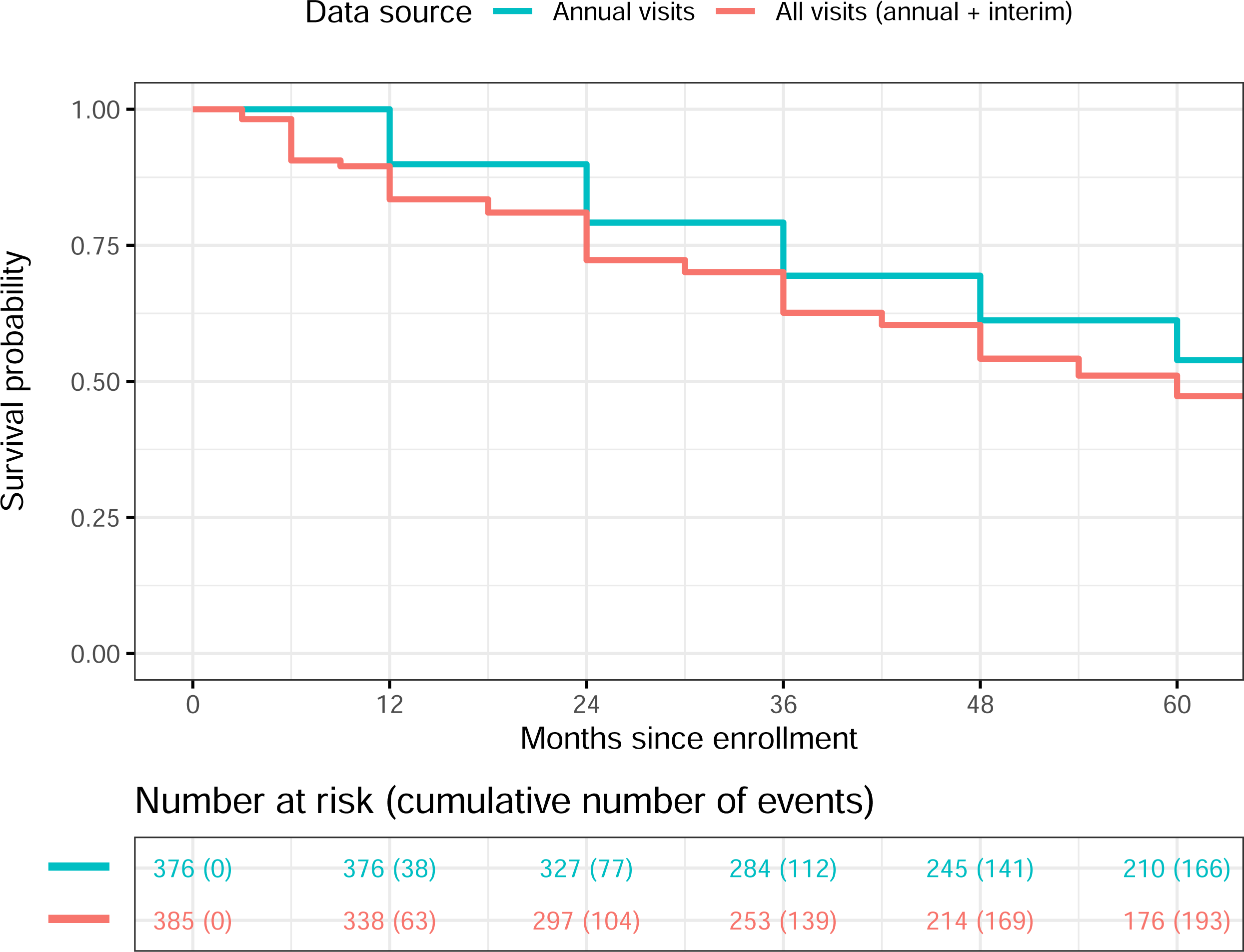
Kaplan-Meier curves of progression-free survival, as defined by reaching any progression milestone, based on data collected at 12, 24, 36, 48, and 60 months (blue) versus 3, 6, 9, 12, 18, 24, 30, 36, 42, 48, 54, and 60 months (red). Each curve reflects de novo PD participants who were milestone-free at baseline and completed at least one of the corresponding follow-up visits.

Table 2 summarizes the contribution of each individual domain and milestone to the composite endpoint, i.e., how frequently they coincided with the *initial* event for a participant. Milestones within the cognitive domain (met by 14.1% of participants based on annual data only vs. 14.3% based on all available data) and functional dependence domain (12.0% vs. 14.5%) were reached first most frequently in this cohort. Collectively, milestones within the autonomic dysfunction (10.9% vs. 10.1%) and walking and balance (6.6% vs. 8.1%) domains were the next most prevalent. Across both data sources, the individual milestone met most often was a S&E below 80.

**Table 2.**
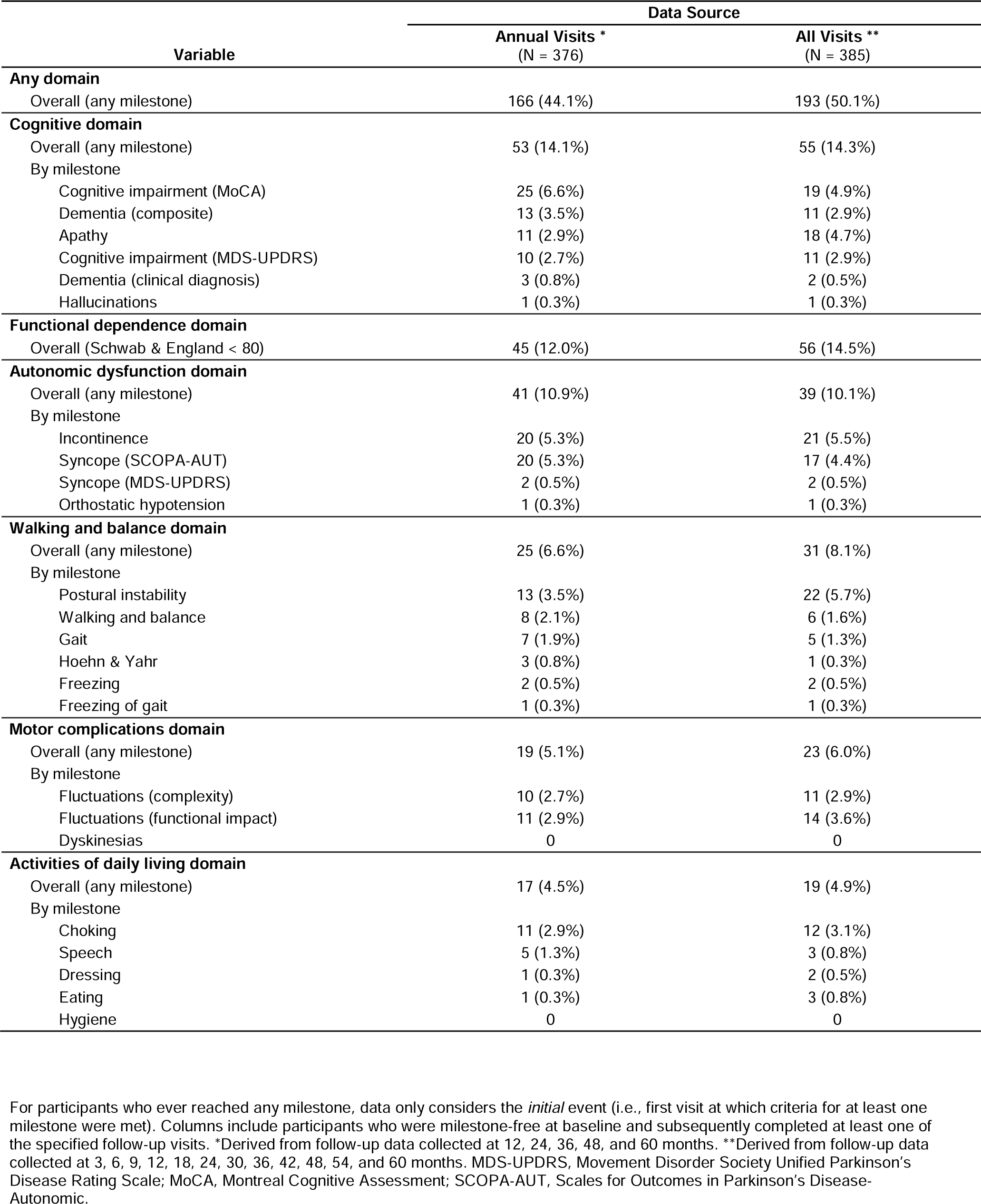
Proportion of PPMI de novo PD cohort that met progression milestone criteria at the time of first event or last follow-up

| Variable | Data Source |  |
| --- | --- | --- |
|  | Annual Visits *<br>(N = 376) | All Visits **<br>(N = 385) |
| **Any domain** |  |  |
| Overall (any milestone) | 166 (44.1%) | 193 (50.1%) |
| **Cognitive domain** |  |  |
| Overall (any milestone) | 53 (14.1%) | 55 (14.3%) |
| By milestone |  |  |
| Cognitive impairment (MoCA) | 25 (6.6%) | 19 (4.9%) |
| Dementia (composite) | 13 (3.5%) | 11 (2.9%) |
| Apathy | 11 (2.9%) | 18 (4.7%) |
| Cognitive impairment (MDS-UPDRS) | 10 (2.7%) | 11 (2.9%) |
| Dementia (clinical diagnosis) | 3 (0.8%) | 2 (0.5%) |
| Hallucinations | 1 (0.3%) | 1 (0.3%) |
| **Functional dependence domain** |  |  |
| Overall (Schwab & England < 80) | 45 (12.0%) | 56 (14.5%) |
| **Autonomic dysfunction domain** |  |  |
| Overall (any milestone) | 41 (10.9%) | 39 (10.1%) |
| By milestone |  |  |
| Incontinence | 20 (5.3%) | 21 (5.5%) |
| Syncope (SCOPA-AUT) | 20 (5.3%) | 17 (4.4%) |
| Syncope (MDS-UPDRS) | 2 (0.5%) | 2 (0.5%) |
| Orthostatic hypotension | 1 (0.3%) | 1 (0.3%) |
| **Walking and balance domain** |  |  |
| Overall (any milestone) | 25 (6.6%) | 31 (8.1%) |
| By milestone |  |  |
| Postural instability | 13 (3.5%) | 22 (5.7%) |
| Walking and balance | 8 (2.1%) | 6 (1.6%) |
| Gait | 7 (1.9%) | 5 (1.3%) |
| Hoehn & Yahr | 3 (0.8%) | 1 (0.3%) |
| Freezing | 2 (0.5%) | 2 (0.5%) |
| Freezing of gait | 1 (0.3%) | 1 (0.3%) |
| **Motor complications domain** |  |  |
| Overall (any milestone) | 19 (5.1%) | 23 (6.0%) |
| By milestone |  |  |
| Fluctuations (complexity) | 10 (2.7%) | 11 (2.9%) |
| Fluctuations (functional impact) | 11 (2.9%) | 14 (3.6%) |
| Dyskinesias | 0 | 0 |
| **Activities of daily living domain** |  |  |
| Overall (any milestone) | 17 (4.5%) | 19 (4.9%) |
| By milestone |  |  |
| Choking | 11 (2.9%) | 12 (3.1%) |
| Speech | 5 (1.3%) | 3 (0.8%) |
| Dressing | 1 (0.3%) | 2 (0.5%) |
| Eating | 1 (0.3%) | 3 (0.8%) |
| Hygiene | 0 | 0 |

For participants who ever reached any milestone, data only considers the *initial* event (i.e., first visit at which criteria for at least one milestone were met). Columns include participants who were milestone-free at baseline and subsequently completed at least one of the specified follow-up visits. \*Derived from follow-up data collected at 12, 24, 36, 48, and 60 months. \*\*Derived from follow-up data collected at 3, 6, 9, 12, 18, 24, 30, 36, 42, 48, 54, and 60 months. MDS-UPDRS, Movement Disorder Society Unified Parkinson's Disease Rating Scale; MoCA, Montreal Cognitive Assessment; SCOPA-AUT, Scales for Outcomes in Parkinson's Disease-Autonomic.

Notably, at the time of the first milestone, most participants met milestone criteria within a single domain only. Among participants who progressed at an annual follow-up, only 26/166 (15.7%) did so across multiple domains concurrently, with 20 reaching milestones within two domains, four within three domains, and two within four domains (data not shown). Among these multi-domain progressors, it was most common for one of the domains to be functional dependence (18/26; 69%), followed by walking and balance (12/26; 46%). By contrast, milestones within the cognition and autonomic dysfunction domains were comparatively likely to occur in isolation, with 43/53 cognitive (81%) and 33/41 (80%) autonomic progressors experiencing an event within a single domain (data not shown).

Descriptive analyses also evaluated the frequency of each milestone in isolation, i.e., if they *ever* occurred regardless of whether a different one occurred first (Supplementary Table 2). Based on data collected at all study visits, 89 participants (23.1%) ever met the functional dependence milestone, 82 (21.3%) ever reached at least one cognitive milestone, and an appreciable number ever reached one or more components of the autonomic dysfunction (16.9%), walking and balance (14.3%), activities of daily living (13.5%), and motor complications (12.5%) domains. Relative to other components of the composite endpoint, those in the activities of daily living domain (choking, speech, dressing, eating, hygiene) were least likely to coincide with the *initial* event; of the 52 participants who ever reached one of these milestones, only 19 did so at their first event (see Table 1).

Table 3 summarizes the analysis of baseline predictors, which modeled time-to-progression based on *annual* milestone assessments only. After adjustment for sex and PD medication use, the final multivariable model included three predictors with positive associations (age, MDS-UPDRS total score, GDS-15 score) and two predictors with negative associations (DAT-SPECT mean striatum SBR, CSF total α-syn). That is, *higher* baseline age, MDS-UPDRS total scores, and GDS-15 scores, as well as *lower* baseline DAT-SPECT binding and CSF total α-syn, predicted faster time to progression. Of note, initiation of symptomatic treatment was not significantly associated with time to progression (*p*=0.1639). Also, a sensitivity analysis excluding participants with CSF hemoglobin above 200 ng/mL (a factor that can potentially impact the results for CSF total α-syn) from the final multivariable model yielded the same conclusions, with the effect of CSF total α-syn strengthening (hazard ratio [95% CI] per decile decrease = 1.12 [1.05, 1.19]; *p* = 0.0004).

**Table 3.** Association between baseline features and time to reaching a progression milestone in PPMI de novo PD cohort

| Baseline Predictor | Screening Analysis |  | Multivariable Analysis |  |
| --- | --- | --- | --- | --- |
|  | HR (90% CI) | P Value | HR (95% CI) | P Value |
| **Demographics** |  |  |  |  |
| Age (per 5-year increase) | 1.25 (1.16, 1.35) | <.0001 | 1.25 (1.13, 1.38) | <.0001 |
| Sex (male vs female) | 0.96 (0.73, 1.26) | 0.8074 | 0.80 (0.58, 1.12) | 0.1996 |
| Clinical site (non-US vs US) | — | 0.7010 | Not Included |  |
| Body mass index | — | 0.6006 | Not Included |  |
| Orthostatic systolic blood pressure change | — | 0.9589 | Not Included |  |
| **Motor features and overall disability** |  |  |  |  |
| Disease duration | — | 0.2087 | Not Included |  |
| MDS-UPDRS total score (per 5-unit increase) | 1.23 (1.18, 1.30) | <.0001 | 1.18 (1.11, 1.26) | <.0001 |
| Hoehn & Yahr stage (stage 2-3 vs stage 1) | 1.52 (1.17, 1.98) | 0.0084 | Not Included |  |
| PIGD score (per 0.1-unit decrease) | 1.19 (1.13, 1.25) | <.0001 | Not Included |  |
| Tremor score | — | 0.1286 | Not Included |  |
| Schwab & England (per 10-unit decrease) | 1.49 (1.20, 1.85) | 0.0024 | — | N.S. |
| PD medication use (yes vs no) * | 1.06 (0.71, 1.60) | 0.8075 | 0.69 (0.41, 1.16) | 0.1639 |
| **Non-Motor Features** |  |  |  |  |
| MoCA (per 1-unit decrease) | 1.10 (1.04, 1.17) | 0.0048 | — | N.S. |
| SCOPA-AUT | 1.08 (1.06, 1.10) | <.0001 | — | N.S. |
| UPSIT (anosmia vs other) | — | 0.1232 | Not Included |  |
| UPSIT raw score (per 1-unit decrease) | — | 0.3262 | Not Included |  |
| Epworth Sleepiness Scale | 1.08 (1.04, 1.12) | 0.0003 | — | N.S. |
| REM Sleep Behavior Disorder Screening Questionnaire | 1.11 (1.06, 1.16) | <.0001 | — | N.S. |
| Geriatric Depression Scale (15-item) | 1.10 (1.05, 1.15) | 0.0004 | 1.07 (1.01, 1.14) | 0.0341 |
| State-Trait Anxiety Inventory total score | 1.01 (1.00, 1.02) | 0.0120 | — | N.S. |
| **Biomarkers** |  |  |  |  |
| DAT-SPECT mean striatum SBR (per 0.1-unit decrease) | 1.11 (1.08, 1.15) | <.0001 | 1.07 (1.02, 1.12) | 0.0043 |
| DAT-SPECT mean putamen SBR (per 0.1-unit decrease) | 1.11 (1.06, 1.17) | 0.0006 | Not Included |  |
| CSF A$\beta_{1-42}$ (per decile decrease) | 1.08 (1.03, 1.13) | 0.0055 | Not Included | |
| CSF *t*-tau | — | 0.1965 | Not Included |  |
| CSF *p*-tau | — | 0.2705 | Not Included |  |
| CSF *t*-tau/A$\beta_{1-42}$ (per decile increase) | 1.07 (1.03, 1.12) | 0.0106 | — | N.S. |
| CSF *t*-tau/A$\beta_{1-42}$ (>0.222 vs <0.222) | 1.47 (1.10, 1.98) | 0.0307 | Not Included | |
| CSF α-synuclein (per decile decrease) | 1.07 (1.03, 1.12) | 0.0097 | 1.09 (1.03, 1.15) | 0.0030 |
| CSF α-synuclein [Hb < 200] (per decile decrease) | 1.10 (1.05, 1.16) | 0.0011 | Not Included | |
| Serum urate | — | 0.6648 | Not Included |  |

Reflects data collected at the first five *annual* follow-up visits only. Multivariable analyses forced sex and initiation of PD medication into the final model. Unless otherwise indicated, hazard ratios for continuous variables were derived in terms of a 1-unit increase.
\*Time-dependent covariate. CSF, cerebrospinal fluid; DAT-SPECT, dopamine transporter single photon emission computed tomography; Hb, hemoglobin; HR, hazard ratio; MDS-UPDRS, Movement Disorder Society Unified Parkinson's Disease Rating Scale; MoCA, Montreal Cognitive Assessment; PIGD, postural instability/gait difficulty; SBR, specific binding ratio; SCOPA-AUT, Scales for Outcomes in Parkinson's Disease-Autonomic; UPSIT, University of Pennsylvania Smell Identification Test.

Additional analyses evaluated the stability of the milestone-based approach at subsequent annual visits (Supplementary Table 3). Of the 166 participants who met criteria for the primary endpoint across the first five annual visits, 14 withdrew without completing an additional annual follow-up (note: 7/14 completed an *interim* visit prior to withdrawal and all seven continued to meet milestone criteria). Among those who remained active in the study, 125/152 (82%) continued to meet criteria for any milestone at one or more subsequent annual visits. Further analyses indicated that participants fell into three broad categories. First, 84 (55%) demonstrated persistent dysfunction, continuing to meet criteria for any milestone at the *next* annual follow-up. Second, 41 (27%) temporarily reverted, not meeting criteria 1 year later but doing so eventually; the period of reversion ranged from 2-5 years, with the majority recurring either 2 years (22/41) or 3 years (12/41) later (data not shown). Third, 27 (18%) permanently reverted, not meeting criteria for any milestone at any subsequent follow-up. Compared to temporary reverters, permanent reverters completed fewer subsequent visits (mean: 3.1 vs. 4.7) and were more likely to complete ≤ 2 additional follow-ups (44% vs. 5%; χ^2^ [1, N = 68] = 15.6, *p* < .0001).

Supplementary Table 3 also presents subgroup analyses summarizing—separately by domain—how often participants met milestone criteria within the *same* domain at the *next* visit, within *any* domain at the *next* visit, and within *any* domain at *any* subsequent visit. Depending on the domain, persistent dysfunction at the next visit ranged from 29-50% if restricted to milestones within the same domain vs. 53-86% if expanded to any milestone from any domain. Rates of permanent reversion (i.e., not meeting criteria within any domain at any subsequent visit) were elevated among individuals whose initial event included milestones within the autonomic dysfunction (22.5%) or motor complications (29.4%) domains.

### Sample Size Calculations

Table 4 presents sample size calculations, based on the survival function estimates depicted in Figure 1, for a two-arm trial targeting 80% power. Estimates vary depending on the source of survival estimates (annual vs. all visits); proposed study length; and, particularly, the assumed treatment effect. For instance, based on the rate of clinically meaningful outcomes we observed in our data, a three-year study assuming a 50% reduction in the hazard ratio would be powered at 80% with approximately 125-150 participants per arm. Alternatively, a three-year study assuming a more modest reduction in the hazard ratio (25%) would likely require at least 600 participants per arm to achieve 80% power.

**Table 4.** Total sample size calculations for a hypothetical two-arm trial using a milestone-based composite endpoint 2-Year Study 3-Year Study

| Data Source | 2-Year Study |  | 3-Year Study |  |
| --- | --- | --- | --- | --- |
|  | HR = 0.50 | HR = 0.75 | HR = 0.50 | HR = 0.75 |
| Annual Visits * | 436 | 2078 | 292 | 1404 |
| All Visits ** | 324 | 1552 | 236 | 1142 |

Hazard ratios of 0.50 and 0.75 denote an assumed reduction in the hazard ratio of 50% and 25%, respectively, among the experimental arm relative to the comparison arm. All calculations specified 80% target power for a two-sided log-rank test at $\alpha = 0.05$ and all estimates reflect the *total* sample size required across two arms. \*Control survival curve estimated based on progression milestone data collected at 12, 24, and 36 months. \*\*Control survival curve estimated based on progression milestone data collected at 3, 6, 9, 12, 18, 24, 30, and 36 months. HR, hazard ratio.

## DISCUSSION

The results of this study show that a set of clinically meaningful milestones derived from widely used assessment scales may have utility as a progression outcome in an early PD cohort. Participants in the PPMI *de novo* PD cohort were recently diagnosed and untreated at the time of entry, and then followed quarterly for one year and semiannually for four years thereafter. Half of this cohort reached at least one milestone during this clinical follow-up period, with over a quarter doing so within two years. The most frequently reached milestones included loss of functional independence, indicators of cognitive impairment (in particular, a MoCA score below 21), measures of dysautonomia (urinary incontinence and syncope), and postural instability. The milestone definitions chosen for this study were intended to reflect more severe forms of a given problem in order to mitigate uncertainty regarding their functional relevance. Importantly, the emergence of milestones was largely independent of whether symptomatic therapy had been initiated. Moreover, the composite endpoint appeared to be relatively stable; among participants who reached at least one milestone and remained active in the study, less than 20% permanently reverted to being “milestone-free” at all subsequent visits. These features support the applicability of a milestone-based outcome measure to assess disease progression in early and middle stage PD.

Multivariable analysis indicated that baseline predictors of faster time to reaching a milestone included advanced age, greater MDS-UPDRS total scores, lower DAT-SPECT striatal binding, lower CSF total α-syn, and higher GDS-15 depression scores. Several of these baseline characteristics—including age, lower DAT binding, and greater motor impairment— have been reported to have poor prognosis in prior studies [29-31], which provides collateral support for our approach. The apparent utility of CSF total α-syn to predict reaching a clinically relevant milestone is especially interesting considering current literature demonstrating that PD is associated with a small but significant decrease in CSF total α-syn concentrations relative to healthy controls [20, 32-34]. These predictors of risk for reaching a milestone suggest enrichment strategies to make clinical trials more efficient by building risk factors into trial entry criteria.

A milestone-based outcome measure offers a degree of adaptability that more conventional methods may lack. For instance, if a participant dropped out early but reached a clinical milestone prior to study withdrawal, this metric of progression would be fully captured in a milestone-based time-to-event model. Because the milestones derived from MDS-UPDRS part III items (measuring gait, freezing of gait, postural stability, and speech) are defined using ON *or* OFF assessment scores, these components can still be evaluated if either the OFF or ON assessment could not be completed. Because of these properties, our results suggest an approach to testing disease-modifying therapies that may not be affected by symptomatic treatment and could be implemented in more naturalistic settings. Specifically, a milestone-based composite endpoint could be considered for trials evaluating novel therapeutics for PD as an add-on to, rather than instead of, standard symptomatic therapy.

Our sample size estimates, which are meant to illustrate the conceptual feasibility of this framework, indicate that the number of participants required for a trial using a milestone-based approach over two to three years would be comparable to a trial of untreated patients using change in MDS-UPDRS over one year of follow-up as the primary outcome measure. This additional follow-up would potentially be balanced by faster recruitment and greater generalizability. By allowing treatment at any point, a milestone-based approach would be both easier to recruit for and more consistent with typical care of PD patients. Furthermore, a milestone-based primary outcome is intrinsically clinically meaningful, while changes in a rating scale could be viewed as an intermediate clinical outcome, without intrinsic meaningfulness [35].

Outcomes measures comprised by a composite of clinically relevant events have been applied in other areas of medicine—including cancer [36], cardiology [37], nephrology [38], and stroke [39]—and have been acceptable to regulators [40]. Milestone-based or composite outcomes have been employed before in PD therapeutics, as well. The Deprenyl and tocopherol antioxidative therapy of parkinsonism (DATATOP) trial [1] defined its primary outcome by a clinically relevant milestone, i.e., the need for dopaminergic therapy. This is similar to our approach but used a single rather than composite outcome. Although a landmark trial, the DATATOP study has been criticized because the outcome was sensitive to the symptomatic effect of selegiline [41]. In this analysis, we focused to select outcomes that would not be substantially influenced by treatment. In addition, we included initiation of symptomatic treatment as a time-dependent covariate in our analyses to control for its effect. The NET-PD study of creatine (LS-1) [42] provides another relevant precedent for our analysis. The LS-1 study used a global statistical test (GST) composed of the modified S&E, Symbol Digit Modalities Test, Parkinson’s Disease Questionnaire (PDQ-39) [43], selected items from the UPDRS and the Modified Rankin Scale [44]. Like our measure, this outcome is composed of clinically meaningful components. Unlike our simple composite measure, scores on the GST did not lend themselves to intuitive clinical interpretation. Thus, our framework for a composite of clinically meaningful outcomes may represent a potential advance over existing metrics in terms of robustness in the setting of symptomatic treatment and clinical interpretability. Other observational cohort studies have included milestone-based or composite outcomes in their analyses. This includes the CamPaIGN study, which examined the “irreversible” milestones of postural instability (Hoehn & Yahr stage 3), dementia, and death [45]; and the Norwegian ParkWest study, which evaluated the “advanced PD” milestones of visual hallucinations, recurrent falls, dementia, and nursing home placement [46]. Other milestones reported in the literature include severe dysphagia, autonomic dysfunction (e.g., orthostatic hypotension), and unintelligible speech [47]. Our study extends the results of those analyses by including additional clinical milestones and more intensive biomarker assessments which potentially make our results more relevant to implementation in therapeutic research.

Our results must be considered in light of several limitations. First, a multidimensional composite may not be appropriate for interventions that are intended to impact only certain contributors to PD disability. Per FDA guidance, composite endpoints should be chosen with an expectation that a given intervention will “have a favorable effect on all the components” [40]. It is possible that the pathophysiological mechanisms underlying the various clinical domains described herein (e.g., motor vs. cognitive vs. autonomic) are too different to expect that a single intervention could favorably affect all of them. However, given that the natural history of PD progression is multifaceted, a clinical endpoint that encompasses both motor and non-motor milestones may be the most appropriate approach to assessing interventions intended to slow overall disease progression [25].

Another limitation is the sheer size of a 25-item composite. For context, a systematic review of 140 cardiovascular trials with a primary composite endpoint published between 2011 and 2016 found that 83% used endpoints with ≤ 4 components whereas only 6% included ≥ 6 components [37]. The FDA recommends choosing composite endpoints with components of “reasonably similar”—and not “substantially different”—clinical importance, a standard that is harder to establish with more components and one to which our composite may not sufficiently adhere [40]. For instance, we report cognitive milestones defined by apathy and hallucinations. Although both symptoms are reported to predict cognitive impairment in PD [29, 48], they are proxy measures and may not clear the bar of being “reasonably similar” to other milestones (e.g., a site investigator diagnosis of dementia). Our data are meant to illustrate the usefulness of the concept of a milestone-based outcome for PD trials. Future directions could include efforts, such as factor analysis, to test our domain grouping system and simplify the composite by removing redundancy and components that contribute minimally to the overall endpoint.

Third, the criteria for our composite endpoint are satisfied by the occurrence of a single rater-dependent event recorded at a single time point, an approach that prioritizes sensitivity over specificity and raises important questions about reliability. We considered an alternative strategy requiring that milestones be evident at *consecutive* visits. However, this made the endpoint less efficient, particularly if participants meeting criteria at baseline were excluded (in which case the endpoint could not be met until the second follow-up visit); and was insensitive to participants who met criteria at a single visit and withdrew before their next visit (due to worsening parkinsonism). Ultimately, we chose a first occurrence strategy, concluding that experiencing something sufficiently severe for the first time represents an important clinical event even if it is not reported at the next visit. Moreover, since cutoffs were made at severe manifestations of each clinical feature, we could envisage medication changes and other therapeutic maneuvers that could temporarily reduce the severity of such problems, which then recur after a hiatus. That said, nearly 20% of participants in our sample who ever reached a milestone did not recur at any subsequent visit. We acknowledge that this is not an insignificant number and that efforts to mitigate such occurrences are warranted. The domain-level analyses reported herein (Supplementary Table 3) suggest that milestones within certain domains (e.g., autonomic dysfunction, motor complications) may be less stable than others and future analyses evaluating the stability of each individual milestone are being planned.

Another important limitation to our study is the lack of Patient and Public Involvement and Engagement (PPIE). Milestones were carefully chosen by a panel of clinical experts and anchored largely to MDS-UPDRS items, which were developed with extensive input from patient focus groups [12]. However, for a milestone-based composite measure to be considered as the primary outcome in a therapeutic trial, greater PPIE would be essential. One possibility would be to survey PD patients and care partners on the relative “clinical importance” of the milestones reported herein and elsewhere in the literature [45-47]. Our composite also lacked a global quality of life measure, such as the PDQ-39, and other patient-reported outcomes (PROs). Additional PROs as well as objective digital measures have been added to the PPMI battery and could be areas of future research.

Other key limitations of our study that warrant further investigation include its exploratory nature (e.g., no adjustment for multiple comparisons) and absence of external validation. Importantly, efforts are underway to validate this milestone-based endpoint in other early PD cohorts, including the STEADY-PD III [49] and SURE-PD3 [50] trial cohorts and their extension in AT-HOME PD [51]. Like PPMI, these studies included participants with early-stage PD who were not on levodopa or dopamine agonists at enrollment. Furthermore, they are comparable in mean age (PPMI = 61.5; STEADY-PD III = 62; SURE-PD3 = 63) and, in the case of SURE-PD3, were similarly enriched for evidence of dopaminergic deficit at screening. Notably, however, these cohorts are considerably younger and far less treated than other PD populations, such as incident PD cases enrolled in the population-based CamPaIGN (mean age: 70.6) and PINE (mean age: 72.5) cohorts [52, 53]. As such, some important considerations will be whether these findings are generalizable to future studies that enroll older and more treated cohorts and which segment of PD patients would be appropriate for a clinical trial that implemented a milestone-based outcome measure.

Also, since milestones were only evaluated at pre-scheduled visits, it only known that criteria became evident at some point during the interval between one visit and the next. However, our analysis used the approach, commonly applied in practice, of assuming that event times were either observed exactly at the *end* of said interval (imputed from the visit date on which the milestone was first recorded) or right censored, and then applying standard time-to-event methods (i.e., Kaplan-Meier and Cox regression estimates). Planned validation efforts will apply methods tailored specifically to interval-censored data [54, 55]. More generally, the use of a time-to-first-event analysis approach may be inefficient because it ignores additional information (e.g., events at subsequent visits, total number of milestones/domains reached). Alternative approaches that may increase study power include recurrent event models [56, 57] or repeated-measures analysis of an ordinal or continuous “score” reflecting the sum of multiple milestones/domains.

These methodological limitations are balanced by important strengths. We conducted our study in the context of the PPMI study which employs rigorous, standardized data collection of motor, non-motor and biomarker assessments in the context of an international, multicenter cohort with long-term follow-up [18, 23, 58]. In summary, the results of this study show that clinically meaningful milestones occur frequently within five years of follow-up of patients recruited with early, untreated PD, and are significantly associated with baseline demographic characteristics, clinical features, and objective biomarkers. These findings support the viability of using a milestone-based outcome measure in observational and biomarker verification studies. Our results also have several important implications for clinical trial design. First, stratification based on baseline markers may reduce variability in progression in clinical trial cohorts, thus making trials more efficient. Second and importantly, a composite measure based on the milestones we evaluated could become a primary outcome in PD disease modification trials. Additional follow-up and analysis of PPMI data will address limitations in our study, produce further validation and refine a framework for efficient trials of potentially disease-modifying therapeutics.

### Supporting information

Supplementary Table 1

Supplementary Table 2

Supplementary Table 3

Supplementary Figure 1

## Data Availability

All data produced are available online at https://www.ppmi-info.org/access-data-specimens/download-data.

^§^ Parkinson’s Progression Markers Initiative Authors

## Executive Steering Committee

Kenneth Marek, MD^1^ (Principal Investigator); Caroline Tanner, MD, PhD^9^; Tanya Simuni, MD^3^; Andrew Siderowf, MD, MSCE^12^; Douglas Galasko, MD^27^; Lana Chahine, MD^41^; Christopher Coffey, PhD^4^; Kalpana Merchant, PhD^61^; Kathleen Poston, MD^40^; Roseanne Dobkin, PhD^43^; Tatiana Foroud, PhD^15^; Brit Mollenhauer, MD^8^; Dan Weintraub, MD^12^; Ethan Brown, MD^9^; Karl Kieburtz, MD, MPH^23^

## Steering Committee

Duygu Tosun-Turgut, PhD^9^; Werner Poewe, MD^7^; Susan Bressman, MD^14^; Jan Hamer^15^; Raymond James, RN^22^; Ekemini Riley, PhD^42^; John Seibyl, MD^1^; Leslie Shaw, PhD^12^; David Standaert, MD, PhD^18^; Sneha Mantri, MD, MS^62^; Nabila Dahodwala, MD^12^; Michael Schwarzschild^47^; Connie Marras^45^; Hubert Fernandez, MD^25^; Ira Shoulson, MD^23^; Helen Rowbotham^2^; Lucy Norcliffe-Kaufmann^2^; Paola Casalin^11^; Claudia Trenkwalder, MD^8^

## Michael J. Fox Foundation (Sponsor)

Todd Sherer, PhD; Sohini Chowdhury; Mark Frasier, PhD; Jamie Eberling, PhD; Katie Kopil, PhD; Alyssa O’Grady; James Gibaldi, MSc; Maggie McGuire Kuhl; Leslie Kirsch, EdD

## Study Cores

*Imaging Core*: John Seibyl, MD^1^; Kathleen Poston, MD^40^

*Statistics Core*: Christopher Coffey, PhD^4^; Chelsea Caspell-Garcia, MS^4^; Janel Fedler, PhD^4^; Michael Brumm, MS^4^; Elliot Burghardt, PhD^4^; Hyunkeun Ryan Cho, PhD^4^; Charlie (Seung Ho) Choi, MS^4^; David-Erick Lafontant, MS^4^

*Bioinformatics Core*: Arthur Toga, PhD^10^; Karen Crawford^10^

## Site Investigators

Ruth Schneider, MD^23^; Kelvin Chou, MD^44^; David Russell, MD, PhD^1^; Stewart Factor, DO^16^; Penelope Hogarth, MD^17^; Robert Hauser, MD, MBA^19^; Nabila Dahodwala, MD, MSc^12^; Marie H Saint-Hilaire, MD, FRCPC, FAAN^22^; David Shprecher, DO^24^; Hubert Fernandez, MD^25^; Kathrin Brockmann, MD^26^; Yen Tai, MD, PhD^29^; Paolo Barone, MD, PhD^30^; Stuart Isaacson, MD^31^; Alberto Espay, MD, MSc, FAAN, FANA^32^; Maria Jose Martí, MD, PhD^34^; Eduardo Tolosa MD, PhD^34^; Shu-Ching Hu, MD, PhD^21^; Douglas Galasko, MD^27^; Emile Moukheiber, MD^28^; Jean-Christophe Corvol, MD^39^; Nir Giladi, MD^36^; Javier Ruiz Martinez, MD, PhD^35^; Jan O. Aasly, MD^37^; Leonidas Stefanis, MD, PhD^38^; Karen Marder, MD MPH^39^; Arjun Tarakad, MD^20^; Connie Marras, MD, PhD, FRCP(C)^45^; Tiago Mestre, MD, PhD^46^; Aleksandar Videnovic, MD, MSc^47^; Rajesh Pahwa, MD^48^; Mark Lew, MD^49^; Holly Shill, MD^50^; Amy Amara, MD, PhD^18^; Charles Adler, MD, PhD^51^; Caroline Tanner, MD, PhD^9^; Susan Bressman, MD^14^; Tanya Simuni, MD^3^; Maureen Leehey, MD^52^; Giulietta Riboldi, MD^53^; Nikolaus McFarland, MD, PhD, FAAN^54^; Lana Chahine, MD^41^; Ron Postuma, MD, FRCPC^55^; Brit Mollenhauer, MD^8^; Werner Poewe, MD^7^; Zoltan Mari, MD^56^; Nicola Pavese, MD, PhD^57^; Michele Hu, MD, PhD^58^; Norbert Brüggemann, MD^59^; Christine Klein, MD, FEAN^59^; Bastiaan Bloem, MD, PhD^60^

## Coordinators

Anisha Singh, BS^23^; Angela Stovall, BS^44^; Julie Festa, BA^1^; Lianne Ramia, BS^1^; Katrina Wakeman, BS^17^; Karen Williams, BA, CCRP^3^; Courtney Blair, MA^18^; Krista Specketer, BS^21^; Diana Willeke^8^; Jennifer Mule, BS^25^; Ella Hilt^26^; Shawnees Peacock, BS^27^; Kori Ribb, RN, BSN, CNRN^28^; Susan Ainscough, BA^30^; Lisbeth Pennente, BA^31^; Julia Brown, BS^32^; Christina Gruenwald, BS, CCRP^32^; Barbara Sommerfeld MSN, RN, CNRN^16^; Farah Kausar, PhD^9^; Alicia Garrido, MD^34^; Deborah Raymond, MS, CGC^14^; Ioana Croitoru^35^; Anne Grete Kristiansen^37^; Helen Mejia Santana, MA^39^; Anjana Singh, BS^20^; Danica Nogo, BS^45^; Shawna Reddie, BA^46^; Samantha Murphy, BS^47^; Lauren O’Brien^48^; Ashwini Ramachandran, MSc^12^; Fnu Madhuri, MS^19^; Daniel Freire, MS^49^; Farah Ismail, MBChB^50^; Raymond James, BS, RN^22^; Tom Osgood, BA, CCRP^51^; Heidi Friedeck, BS^3^; Jenny Frisendahl, BS^52^; Ying Liu, MD^52^; Caitlin Romano, BA^53^; Kelly Clark^24^; Kyle Rizer, BA^54^; Stephanie Carvalho^39^; Sherri Mosovsky, MPH^41^; Farah Sulaiman, MPH^55^; Dora Valent, MS^7^; Raquel Lopes, BSN, MS^29^; Michelle Torreliza, AS^56^; Shira Paz, BS^36^; Victoria Kate Foster^57^; Madita Grümmer^59^; Myrthe Burgler, MA^60^; Sabine van Zundert, MS^60^; Christos Koros, MD, PhD^38^; Jamil Razzaque, MS^58^

## Affiliations

^1^ Institute for Neurodegenerative Disorders, New Haven, CT

^2^ 23andMe

^3^ Northwestern University, Chicago, IL

^4^ University of Iowa, Iowa City, IA

^5^ VectivBio AG

^6^ The Michael J. Fox Foundation for Parkinson’s Research, New York, NY

^7^ Innsbruck Medical University, Innsbruck, Austria

^8^ Paracelsus-Elena Klinik, Kassel, Germany

^9^ University of California, San Francisco, CA

^10^ Laboratory of Neuroimaging (LONI), University of Southern California

^11^ BioRep, Milan, Italy

^12^ University of Pennsylvania, Philadelphia, PA

^13^ National Institute on Aging, NIH, Bethesda, MD

^14^ Mount Sinai Beth Israel, New York, NY

^15^ Indiana University, Indianapolis, IN

^16^ Emory University of Medicine, Atlanta, GA

^17^ Oregon Health and Science University, Portland, OR

^18^ University of Alabama at Birmingham, Birmingham, AL

^19^ University of South Florida, Tampa, FL

^20^ Baylor College of Medicine, Houston, TX

^21^ University of Washington, Seattle, WA

^22^ Boston University, Boston, MA

^23^ University of Rochester, Rochester, NY

^24^ Banner Research Institute, Sun City, AZ

^25^ Cleveland Clinic, Cleveland, OH

^26^ University of Tuebingen, Tuebingen, Germany

^27^ University of California, San Diego, CA

^28^ Johns Hopkins University, Baltimore, MD

^29^ Imperial College of London, London, UK

^30^ University of Salerno, Salerno, Italy

^31^ Parkinson’s Disease and Movement Disorders Center, Boca Raton, FL

^32^ University of Cincinnati, Cincinnati, OH

^34^ Hospital Clinic of Barcelona, Barcelona, Spain

^35^ Hospital Universitario Donostia, San Sebastian, Spain

^36^ Tel Aviv Sourasky Medical Center, Tel Aviv, Israel

^37^ St. Olav’s University Hospital, Trondheim, Norway

^38^ National and Kapodistrian University of Athens, Athens, Greece

^39^ Columbia University Irving Medical Center, New York, NY

^40^ Stanford University, Stanford, CA

^41^ University of Pittsburgh, Pittsburgh, PA

^42^ Center for Strategy Philanthropy at Milken Institute, Washington D.C.

^43^ Rutgers University, New Brunswick, NJ

^44^ University of Michigan, Ann Arbor, MI

^45^ Toronto Western Hospital, Toronto, Canada

^46^ The Ottawa Hospital, Ottawa, Canada

^47^ Massachusetts General Hospital, Boston, MA

^48^ University of Kansas Medical Center, Kansas City, KS

^49^ University of Southern California, Los Angeles, CA

^50^ Barrow Neurological Institute, Phoenix, AZ

^51^ Mayo Clinic Arizona, Scottsdale, AZ

^52^ University of Colorado, Aurora, CO

^53^ NYU Langone Medical Center, New York, NY

^54^ University of Florida, Gainesville, FL

^55^ Montreal Neurological Institute and Hospital/McGill, Montreal, QC, Canada

^56^ Cleveland Clinic-Las Vegas Lou Ruvo Center for Brain Health, Las Vegas, NV

^57^ Clinical Ageing Research Unit, Newcastle, UK

^58^ John Radcliffe Hospital Oxford and Oxford University, Oxford, UK

^59^ Universität Lübeck, Luebeck, Germany

^60^ Radboud University, Nijmegen, Netherlands

^61^ TransThera Consulting

^62^ Duke University, Durham, NC

## ACKNOWLEDGEMENTS

PPMI – a public-private partnership – is funded by the Michael J. Fox Foundation for Parkinson’s Research funding partners 4D Pharma, Abbvie, Acurex Therapeutics, Aligning Science Across Parkinson’s, Allergan, Amathus Therapeutics, Avid Radiopharmaceuticals, Bial Biotech, Biogen, BioLegend, Bristol-Myers Squibb, Calico, Celgene, Dacapo Brain Science, Denali, The Edmond J. Safra Foundaiton, GE Healthcare, Genentech, GlaxoSmithKline, Golub Capital, Handl Therapeutics, Insitro, Janssen Neuroscience, Lilly, Lundbeck, Merck, Meso Scale Discovery, Neurocrine Biosciences, Pfizer, Piramal, Prevail, Roche, Sanofi Genzyme, Servier, Takeda, Teva, UCB, Verily, and Voyager Therapeutics.

We are grateful to the Michael J. Fox Foundation for Parkinson’s Research, all our PPMI colleagues, and the many individuals who sacrificed their time and gave of themselves to be participants in this study.

---

Supplementary Fig. 1. Flow chart summarizing which participants from PPMI *de novo* PD cohort were included in the analysis and how many participants were assessed at each study time point.

## REFERENCES

[1] (1993) Effects of tocopherol and deprenyl on the progression of disability in early Parkinson’s disease. N Engl J Med 328, 176–183.

[2] de Lau LM, Schipper CM, Hofman A, Koudstaal PJ, Breteler MM (2005) Prognosis of Parkinson disease: risk of dementia and mortality: the Rotterdam Study. Arch Neurol 62, 1265–1269.

[3] De Pablo-Fernandez E, Tur C, Revesz T, Lees AJ, Holton JL, Warner TT (2017) Association of Autonomic Dysfunction With Disease Progression and Survival in Parkinson Disease. JAMA Neurol 74, 970–976.

[4] Levy G, Tang MX, Louis ED, Cote LJ, Alfaro B, Mejia H, Stern Y, Marder K (2002) The association of incident dementia with mortality in PD. Neurology 59, 1708–1713.

[5] Merola A, Coon EA (2020) Dysautonomia in early Parkinson disease: a window into the determinants of functional disability and an opportunity for early intervention. Clin Auton Res 30, 191–192.

[6] Williams-Gray CH, Foltynie T, Brayne CE, Robbins TW, Barker RA (2007) Evolution of cognitive dysfunction in an incident Parkinson’s disease cohort. Brain 130, 1787–1798.

[7] Hely MA, Reid WG, Adena MA, Halliday GM, Morris JG (2008) The Sydney multicenter study of Parkinson’s disease: the inevitability of dementia at 20 years. Mov Disord 23, 837–844.

[8] Huang P, Goetz CG, Woolson RF, Tilley B, Kerr D, Palesch Y, Elm J, Ravina B, Bergmann KJ, Kieburtz K, Parkinson Study G (2009) Using global statistical tests in long-term Parkinson’s disease clinical trials. Mov Disord 24, 1732–1739.

[9] Simuni T, Brumm MC, Uribe L, Caspell-Garcia C, Coffey CS, Siderowf A, Alcalay RN, Trojanowski JQ, Shaw LM, Seibyl J, Singleton A, Toga AW, Galasko D, Foroud T, Nudelman K, Tosun-Turgut D, Poston K, Weintraub D, Mollenhauer B, Tanner CM, Kieburtz K, Chahine LM, Reimer A, Hutten S, Bressman S, Marek K, Parkinson’s Progression Markers Initiative I (2020) Clinical and Dopamine Transporter Imaging Characteristics of Leucine Rich Repeat Kinase 2 (LRRK2) and Glucosylceramidase Beta (GBA) Parkinson’s Disease Participants in the Parkinson’s Progression Markers Initiative: A Cross-Sectional Study. Mov Disord 35, 833–844.

[10] Weintraub D, Caspell-Garcia C, Simuni T, Cho HR, Coffey CS, Aarsland D, Alcalay RN, Barrett MJ, Chahine LM, Eberling J, Espay AJ, Hamilton J, Hawkins KA, Leverenz J, Litvan I, Richard I, Rosenthal LS, Siderowf A, York M, Parkinson’s Progression Markers I (2020) Neuropsychiatric symptoms and cognitive abilities over the initial quinquennium of Parkinson disease. Ann Clin Transl Neurol 7, 449–461.

[11] Fahn S, Elton R (1987) UPDRS program members. Unified Parkinsons disease rating scale. Recent developments in Parkinson’s disease 2, 153–163.

[12] Goetz CG, Tilley BC, Shaftman SR, Stebbins GT, Fahn S, Martinez-Martin P, Poewe W, Sampaio C, Stern MB, Dodel R, Dubois B, Holloway R, Jankovic J, Kulisevsky J, Lang AE, Lees A, Leurgans S, LeWitt PA, Nyenhuis D, Olanow CW, Rascol O, Schrag A, Teresi JA, van Hilten JJ, LaPelle N (2008) Movement Disorder Society-sponsored revision of the Unified Parkinson’s Disease Rating Scale (MDS-UPDRS): scale presentation and clinimetric testing results. Mov Disord 23, 2129–2170.

[13] Martinez-Martin P, Rodriguez-Blazquez C, Alvarez-Sanchez M, Arakaki T, Bergareche-Yarza A, Chade A, Garretto N, Gershanik O, Kurtis MM, Martinez-Castrillo JC, Mendoza-Rodriguez A, Moore HP, Rodriguez-Violante M, Singer C, Tilley BC, Huang J, Stebbins GT, Goetz CG (2013) Expanded and independent validation of the Movement Disorder Society-Unified Parkinson’s Disease Rating Scale (MDS-UPDRS). J Neurol 260, 228–236.

[14] Movement Disorder Society Task Force on Rating Scales for Parkinson’s D (2003) The Unified Parkinson’s Disease Rating Scale (UPDRS): status and recommendations. Mov Disord 18, 738–750.

[15] Schrag A, Spottke A, Quinn NP, Dodel R (2009) Comparative responsiveness of Parkinson’s disease scales to change over time. Mov Disord 24, 813–818.

[16] Braunwald E, Cannon CP, McCabe CH (1993) Use of composite endpoints in thrombolysis trials of acute myocardial infarction. Am J Cardiol 72, 3G–12G.

[17] (2011) The Parkinson Progression Marker Initiative (PPMI). Prog Neurobiol 95, 629–635.

[18] Marek K, Chowdhury S, Siderowf A, Lasch S, Coffey CS, Caspell-Garcia C, Simuni T, Jennings D, Tanner CM, Trojanowski JQ, Shaw LM, Seibyl J, Schuff N, Singleton A, Kieburtz K, Toga AW, Mollenhauer B, Galasko D, Chahine LM, Weintraub D, Foroud T, Tosun-Turgut D, Poston K, Arnedo V, Frasier M, Sherer T (2018) The Parkinson’s progression markers initiative (PPMI) - establishing a PD biomarker cohort. Ann Clin Transl Neurol 5, 1460–1477.

[19] Stebbins GT, Goetz CG, Burn DJ, Jankovic J, Khoo TK, Tilley BC (2013) How to identify tremor dominant and postural instability/gait difficulty groups with the movement disorder society unified Parkinson’s disease rating scale: comparison with the unified Parkinson’s disease rating scale. Mov Disord 28, 668–670.

[20] Mollenhauer B, Caspell-Garcia CJ, Coffey CS, Taylor P, Singleton A, Shaw LM, Trojanowski JQ, Frasier M, Simuni T, Iranzo A, Oertel W, Siderowf A, Weintraub D, Seibyl J, Toga AW, Tanner CM, Kieburtz K, Chahine LM, Marek K, Galasko D (2019) Longitudinal analyses of cerebrospinal fluid alpha-Synuclein in prodromal and early Parkinson’s disease. Mov Disord 34, 1354–1364.

[21] Irwin DJ, Fedler J, Coffey CS, Caspell-Garcia C, Kang JH, Simuni T, Foroud T, Toga AW, Tanner CM, Kieburtz K, Chahine LM, Reimer A, Hutten S, Weintraub D, Mollenhauer B, Galasko DR, Siderowf A, Marek K, Trojanowski JQ, Shaw LM, Parkinson’s Progression Marker I (2020) Evolution of Alzheimer’s Disease Cerebrospinal Fluid Biomarkers in Early Parkinson’s Disease. Ann Neurol 88, 574–587.

[22] Simuni T, Long JD, Caspell-Garcia C, Coffey CS, Lasch S, Tanner CM, Jennings D, Kieburtz KD, Marek K (2016) Predictors of time to initiation of symptomatic therapy in early Parkinson’s disease. Ann Clin Transl Neurol 3, 482–494.

[23] Weintraub D, Simuni T, Caspell-Garcia C, Coffey C, Lasch S, Siderowf A, Aarsland D, Barone P, Burn D, Chahine LM, Eberling J, Espay AJ, Foster ED, Leverenz JB, Litvan I, Richard I, Troyer MD, Hawkins KA (2015) Cognitive performance and neuropsychiatric symptoms in early, untreated Parkinson’s disease. Mov Disord 30, 919–927.

[24] Emre M, Aarsland D, Brown R, Burn DJ, Duyckaerts C, Mizuno Y, Broe GA, Cummings J, Dickson DW, Gauthier S, Goldman J, Goetz C, Korczyn A, Lees A, Levy R, Litvan I, McKeith I, Olanow W, Poewe W, Quinn N, Sampaio C, Tolosa E, Dubois B (2007) Clinical diagnostic criteria for dementia associated with Parkinson’s disease. Mov Disord 22, 1689–1707; quiz 1837.

[25] Poewe W (2006) The natural history of Parkinson’s disease. J Neurol 253 Suppl 7, VII2–6.

[26] Caspell-Garcia C, Simuni T, Tosun-Turgut D, Wu IW, Zhang Y, Nalls M, Singleton A, Shaw LA, Kang JH, Trojanowski JQ, Siderowf A, Coffey C, Lasch S, Aarsland D, Burn D, Chahine LM, Espay AJ, Foster ED, Hawkins KA, Litvan I, Richard I, Weintraub D (2017) Multiple modality biomarker prediction of cognitive impairment in prospectively followed de novo Parkinson disease. PLoS One 12, e0175674.

[27] Kassambara A, Kosinski M, Biecek P, Fabian S (2017) survminer: Drawing Survival Curves using’ggplot2’. R package version 0.3 1.

[28] Mollenhauer B, Batrla R, El-Agnaf O, Galasko DR, Lashuel HA, Merchant KM, Shaw LM, Selkoe DJ, Umek R, Vanderstichele H, Zetterberg H, Zhang J, Caspell-Garcia C, Coffey C, Hutten SJ, Frasier M, Taylor P (2017) A user’s guide for α-synuclein biomarker studies in biological fluids: Perianalytical considerations. Mov Disord 32, 1117–1130.

[29] Aarsland D, Andersen K, Larsen JP, Lolk A, Kragh-Sorensen P (2003) Prevalence and characteristics of dementia in Parkinson disease: an 8-year prospective study. Arch Neurol 60, 387–392.

[30] McDermott MP, Jankovic J, Carter J, Fahn S, Gauthier S, Goetz CG, Golbe LI, Koller W, Lang AE, Olanow CW, et al. (1995) Factors predictive of the need for levodopa therapy in early, untreated Parkinson’s disease. The Parkinson Study Group. Arch Neurol 52, 565–570.

[31] Ravina B, Marek K, Eberly S, Oakes D, Kurlan R, Ascherio A, Beal F, Beck J, Flagg E, Galpern WR, Harman J, Lang AE, Schwarzschild M, Tanner C, Shoulson I (2012) Dopamine transporter imaging is associated with long-term outcomes in Parkinson’s disease. Mov Disord 27, 1392–1397.

[32] Mollenhauer B, Trautmann E, Taylor P, Manninger P, Sixel-Doring F, Ebentheuer J, Trenkwalder C, Schlossmacher MG (2013) Total CSF alpha-synuclein is lower in de novo Parkinson patients than in healthy subjects. Neurosci Lett 532, 44–48.

[33] Hong Z, Shi M, Chung KA, Quinn JF, Peskind ER, Galasko D, Jankovic J, Zabetian CP, Leverenz JB, Baird G, Montine TJ, Hancock AM, Hwang H, Pan C, Bradner J, Kang UJ, Jensen PH, Zhang J (2010) DJ-1 and alpha-synuclein in human cerebrospinal fluid as biomarkers of Parkinson’s disease. Brain 133, 713–726.

[34] Kang JH, Irwin DJ, Chen-Plotkin AS, Siderowf A, Caspell C, Coffey CS, Waligorska T, Taylor P, Pan S, Frasier M, Marek K, Kieburtz K, Jennings D, Simuni T, Tanner CM, Singleton A, Toga AW, Chowdhury S, Mollenhauer B, Trojanowski JQ, Shaw LM, Parkinson’s Progression Markers I (2013) Association of cerebrospinal fluid beta-amyloid 1-42, T-tau, P-tau181, and alpha-synuclein levels with clinical features of drug-naive patients with early Parkinson disease. JAMA Neurol 70, 1277–1287.

[35] (2014) US Department of Health and Human Services: Center for Drug Evaluation and Research (CDER) and Center for Biologics Evaluation and Research (CBER).

[36] Stopeck AT, Lipton A, Body JJ, Steger GG, Tonkin K, de Boer RH, Lichinitser M, Fujiwara Y, Yardley DA, Viniegra M, Fan M, Jiang Q, Dansey R, Jun S, Braun A (2010) Denosumab compared with zoledronic acid for the treatment of bone metastases in patients with advanced breast cancer: a randomized, double-blind study. J Clin Oncol 28, 5132–5139.

[37] Armstrong PW, Westerhout CM (2017) Composite End Points in Clinical Research: A Time for Reappraisal. Circulation 135, 2299–2307.

[38] Brenner BM, Cooper ME, de Zeeuw D, Keane WF, Mitch WE, Parving HH, Remuzzi G, Snapinn SM, Zhang Z, Shahinfar S, Investigators RS (2001) Effects of losartan on renal and cardiovascular outcomes in patients with type 2 diabetes and nephropathy. N Engl J Med 345, 861–869.

[39] National Institute of Neurological D, Stroke rt PASSG (1995) Tissue plasminogen activator for acute ischemic stroke. N Engl J Med 333, 1581–1587.

[40] Food and Drug Administration (2017) Multiple endpoints in clinical trials: guidance for industry.

[41] Landau WM (1990) Clinical neuromythology IX. Pyramid sale in the bucket shop: DATATOP bottoms out. Neurology 40, 1337–1339.

[42] Kieburtz K, Tilley BC, Elm JJ, Babcock D, Hauser R, Ross GW, Augustine AH, Augustine EU, Aminoff MJ, Bodis-Wollner IG, Boyd J, Cambi F, Chou K, Christine CW, Cines M, Dahodwala N, Derwent L, Dewey RB, Jr., Hawthorne K, Houghton DJ, Kamp C, Leehey M, Lew MF, Liang GS, Luo ST, Mari Z, Morgan JC, Parashos S, Pérez A, Petrovitch H, Rajan S, Reichwein S, Roth JT, Schneider JS, Shannon KM, Simon DK, Simuni T, Singer C, Sudarsky L, Tanner CM, Umeh CC, Williams K, Wills AM (2015) Effect of creatine monohydrate on clinical progression in patients with Parkinson disease: a randomized clinical trial. Jama 313, 584–593.

[43] Peto V, Jenkinson C, Fitzpatrick R (1998) PDQ-39: a review of the development, validation and application of a Parkinson’s disease quality of life questionnaire and its associated measures. J Neurol 245 Suppl 1, S10–14.

[44] Elm JJ (2012) Design innovations and baseline findings in a long-term Parkinson’s trial: the National Institute of Neurological Disorders and Stroke Exploratory Trials in Parkinson’s Disease Long-Term Study-1. Mov Disord 27, 1513–1521.

[45] Williams-Gray CH, Mason SL, Evans JR, Foltynie T, Brayne C, Robbins TW, Barker RA (2013) The CamPaIGN study of Parkinson’s disease: 10-year outlook in an incident population-based cohort. J Neurol Neurosurg Psychiatry 84, 1258–1264.

[46] Gonzalez MC, Dalen I, Maple-Grodem J, Tysnes OB, Alves G (2022) Parkinson’s disease clinical milestones and mortality. NPJ Parkinsons Dis 8, 58.

[47] Marsili L, Mahajan A (2022) Clinical milestones in Parkinson’s disease: Past, present, and future. J Neurol Sci 432, 120082.

[48] Martin GP, McDonald KR, Allsop D, Diggle PJ, Leroi I (2020) Apathy as a behavioural marker of cognitive impairment in Parkinson’s disease: a longitudinal analysis. J Neurol 267, 214–227.

[49] Parkinson Study Group S-PDIIII (2020) Isradipine Versus Placebo in Early Parkinson Disease: A Randomized Trial. Ann Intern Med 172, 591–598.

[50] Parkinson Study Group S-PDI, Schwarzschild MA, Ascherio A, Casaceli C, Curhan GC, Fitzgerald R, Kamp C, Lungu C, Macklin EA, Marek K, Mozaffarian D, Oakes D, Rudolph A, Shoulson I, Videnovic A, Scott B, Gauger L, Aldred J, Bixby M, Ciccarello J, Gunzler SA, Henchcliffe C, Brodsky M, Keith K, Hauser RA, Goetz C, LeDoux MS, Hinson V, Kumar R, Espay AJ, Jimenez-Shahed J, Hunter C, Christine C, Daley A, Leehey M, de Marcaida JA, Friedman JH, Hung A, Bwala G, Litvan I, Simon DK, Simuni T, Poon C, Schiess MC, Chou K, Park A, Bhatti D, Peterson C, Criswell SR, Rosenthal L, Durphy J, Shill HA, Mehta SH, Ahmed A, Deik AF, Fang JY, Stover N, Zhang L, Dewey RB, Jr., Gerald A, Boyd JT, Houston E, Suski V, Mosovsky S, Cloud L, Shah BB, Saint-Hilaire M, James R, Zauber SE, Reich S, Shprecher D, Pahwa R, Langhammer A, LaFaver K, LeWitt PA, Kaminski P, Goudreau J, Russell D, Houghton DJ, Laroche A, Thomas K, McGraw M, Mari Z, Serrano C, Blindauer K, Rabin M, Kurlan R, Morgan JC, Soileau M, Ainslie M, Bodis-Wollner I, Schneider RB, Waters C, Ratel AS, Beck CA, Bolger P, Callahan KF, Crotty GF, Klements D, Kostrzebski M, McMahon GM, Pothier L, Waikar SS, Lang A, Mestre T (2021) Effect of Urate-Elevating Inosine on Early Parkinson Disease Progression: The SURE-PD3 Randomized Clinical Trial. JAMA 326, 926–939.

[51] Schneider RB, Omberg L, Macklin EA, Daeschler M, Bataille L, Anthwal S, Myers TL, Baloga E, Duquette S, Snyder P, Amodeo K, Tarolli CG, Adams JL, Callahan KF, Gottesman J, Kopil CM, Lungu C, Ascherio A, Beck JC, Biglan K, Espay AJ, Tanner C, Oakes D, Shoulson I, Novak D, Kayson E, Ray Dorsey E, Mangravite L, Schwarzschild MA, Simuni T, Parkinson Study Group ATHPDI (2021) Design of a virtual longitudinal observational study in Parkinson’s disease (AT-HOME PD). Ann Clin Transl Neurol 8, 308–320.

[52] Foltynie T, Brayne CE, Robbins TW, Barker RA (2004) The cognitive ability of an incident cohort of Parkinson’s patients in the UK. The CamPaIGN study. Brain 127, 550–560.

[53] Caslake R, Taylor K, Scott N, Gordon J, Harris C, Wilde K, Murray A, Counsell C (2013) Age-, gender-, and socioeconomic status-specific incidence of Parkinson’s disease and parkinsonism in northeast Scotland: the PINE study. Parkinsonism Relat Disord 19, 515–521.

[54] Lindsey JC, Ryan LM (1998) Tutorial in biostatistics methods for interval-censored data. Stat Med 17, 219–238.

[55] Gómez G, Calle ML, Oller R, Langohr K (2009) Tutorial on methods for interval-censored data and their implementation in R. Statistical Modelling 9, 259–297.

[56] Amorim LD, Cai J (2015) Modelling recurrent events: a tutorial for analysis in epidemiology. Int J Epidemiol 44, 324–333.

[57] Ozga AK, Rauch G (2022) Weighted composite time to event endpoints with recurrent events: comparison of three analytical approaches. BMC Med Res Methodol 22, 38.

[58] Simuni T, Siderowf A, Lasch S, Coffey CS, Caspell-Garcia C, Jennings D, Tanner CM, Trojanowski JQ, Shaw LM, Seibyl J, Schuff N, Singleton A, Kieburtz K, Toga AW, Mollenhauer B, Galasko D, Chahine LM, Weintraub D, Foroud T, Tosun D, Poston K, Arnedo V, Frasier M, Sherer T, Chowdhury S, Marek K, Parkinson’s Progression Marker I (2018) Longitudinal Change of Clinical and Biological Measures in Early Parkinson’s Disease: Parkinson’s Progression Markers Initiative Cohort. Mov Disord 33, 771–782.

