## Supplementary Table 1 for "Parkinson’s Progression Markers Initiative: A Milestone-Based Strategy to Monitor PD Progression"

**Supplementary Table 1. Baseline demographic and disease characteristics**

|  | **Data Source** | |
| --- | --- | --- |
| **Characteristic** | **Annual Visits** (N = 376) | **All Visits** (N = 385) |
| **Age at enrollment**, mean (SD) | 61.5 (9.8) | 61.5 (9.8) |
| **Sex**, n (%) |  |  |
| Female | 127 (33.8%) | 130 (33.8%) |
| Male | 249 (66.2%) | 255 (66.2%) |
| **Clinical site**, n (%) * |  |  |
| Non-US | 69 (18.4%) | 70 (18.2%) |
| US | 307 (81.6%) | 315 (81.8%) |
| **Race**, n (%) ** |  |  |
| Asian | 8 (2.1%) | 8 (2.1%) |
| Black or African American | 6 (1.6%) | 6 (1.6%) |
| Multiracial | 6 (1.6%) | 6 (1.6%) |
| White | 355 (94.7%) | 364 (94.8%) |
| **Ethnicity**, n (%) |  |  |
| Hispanic or Latino | 5 (1.3%) | 5 (1.3%) |
| Not Hispanic or Latino | 371 (98.7%) | 380 (98.7%) |
| **Years of education**, mean (SD) | 15.6 (2.9) | 15.6 (2.9) |
| **Body mass index (kg/m^2^)**, mean (SD) | 27.2 (4.7) | 27.2 (4.7) |
| **Orthostatic systolic blood pressure change**, mean (SD) | 4.8 (12.4) | 4.8 (12.4) |
| **Disease duration from diagnosis (months)**, mean (SD) | 6.7 (6.5) | 6.6 (6.5) |
| **Family history of PD (first-degree relatives)**, n (%) | 48 (12.8%) | 50 (13.0%) |
| **MDS-UPDRS total score**, mean (SD) | 31.5 (12.8) | 31.5 (12.7) |
| **Hoehn & Yahr stage**, mean (SD) | 1.6 (0.5) | 1.6 (0.5) |
| **Schwab & England**, mean (SD) | 93.5 (5.6) | 93.5 (5.6) |
| **Montreal Cognitive Assessment score**, mean (SD) | 27.2 (2.2) | 27.2 (2.2) |
| **SCOPA-AUT**, mean (SD) | 9.0 (5.5) | 9.0 (5.5) |
| **DAT-SPECT mean striatum SBR**, mean (SD) | 1.40 (0.39) | 1.40 (0.39) |
| **Serum urate (mg/dL)**, mean (SD) | 5.4 (1.3) | 5.4 (1.3) |

Columns include participants who were milestone-free at baseline and subsequently completed at least one annual follow-up visit (i.e., 12, 24, 36, 48, and/or 60 months) or at least one annual or interim follow-up visit (i.e., 3, 6, 9, 12, 18, 24, 30, 36, 42, 48, 54, and 60 months). *Non-US participants were recruited from sites in Australia (3), Austria (7), England (5), Germany (45), and Italy (10). **Multiracial subgroup includes 4 participants who self-identified as American Indian or Alaska Native and White and 2 who self-identified as Asian and White; race is missing for 1 participant. DAT-SPECT, dopamine transporter single photon emission computed tomography; MDS-UPDRS, Movement Disorder Society Unified Parkinson’s Disease Rating Scale; SBR, specific binding ratio; SCOPA-AUT, Scales for Outcomes in Parkinson’s Disease-Autonomic.
