## Supplementary Table 2 for "Parkinson’s Progression Markers Initiative: A Milestone-Based Strategy to Monitor PD Progression"

**Supplementary Table 2. Proportion of PPMI *de novo* PD cohort that *ever* met progression milestone criteria within five years**

|  | **Data Source** | |
| --- | --- | --- |
| **Variable** | **Annual Visits *** (N = 376) | **All Visits **** (N = 385) |
| **Any domain** |  |  |
| Overall (any milestone) | 166 (44.1%) | 193 (50.1%) |
| **Cognitive domain** |  |  |
| Overall (any milestone) | 71 (18.9%) | 82 (21.3%) |
| By milestone |  |  |
| Cognitive impairment (MoCA) | 37 (9.8%) | 37 (9.8%) |
| Dementia (composite) | 25 (6.7%) | 25 (6.7%) |
| Apathy | 20 (5.3%) | 32 (8.3%) |
| Cognitive impairment (MDS-UPDRS) | 26 (6.9%) | 34 (8.8%) |
| Dementia (clinical diagnosis) | 18 (4.9%) | 18 (4.9%) |
| Hallucinations | 6 (1.6%) | 7 (1.8%) |
| **Functional dependence domain** |  |  |
| Overall (Schwab & England < 80) | 72 (19.1%) | 89 (23.1%) |
| **Autonomic dysfunction domain** |  |  |
| Overall (any milestone) | 60 (16.0%) | 65 (16.9%) |
| By milestone |  |  |
| Incontinence | 34 (9.1%) | 36 (9.4%) |
| Syncope (SCOPA-AUT) | 31 (8.2%) | 33 (8.6%) |
| Syncope (MDS-UPDRS) | 4 (1.1%) | 7 (1.8%) |
| Orthostatic hypotension | 5 (1.3%) | 5 (1.3%) |
| **Walking and balance domain** |  |  |
| Overall (any milestone) | 41 (10.9%) | 55 (14.3%) |
| By milestone |  |  |
| Postural instability | 29 (7.8%) | 39 (10.1%) |
| Walking and balance | 22 (5.9%) | 28 (7.3%) |
| Gait | 21 (5.6%) | 25 (6.5%) |
| Hoehn & Yahr | 15 (4.0%) | 16 (4.2%) |
| Freezing | 10 (2.7%) | 14 (3.6%) |
| Freezing of gait | 4 (1.1%) | 5 (1.3%) |
| **Motor complications domain** |  |  |
| Overall (any milestone) | 33 (8.8%) | 48 (12.5%) |
| By milestone |  |  |
| Fluctuations (complexity) | 15 (4.0%) | 22 (5.7%) |
| Fluctuations (functional impact) | 23 (6.1%) | 35 (9.1%) |
| Dyskinesias | 2 (0.5%) | 3 (0.8%) |
| **Activities of daily living domain** |  |  |
| Overall (any milestone) | 29 (7.7%) | 52 (13.5%) |
| By milestone |  |  |
| Choking | 13 (3.5%) | 24 (6.2%) |
| Speech | 11 (2.9%) | 12 (3.1%) |
| Dressing | 9 (2.4%) | 21 (5.5%) |
| Eating | 6 (1.6%) | 13 (3.4%) |
| Hygiene | 4 (1.1%) | 5 (1.3%) |

Data indicates the proportion of participants who ever met criteria for each individual milestone within five years of follow-up irrespective of the order of occurrence (i.e., regardless of whether a different milestone was reached first). Columns include participants who were milestone-free at baseline and subsequently completed at least one of the specified follow-up visits. *Derived from follow-up data collected at 12, 24, 36, 48, and 60 months. **Derived from follow-up data collected at 3, 6, 9, 12, 18, 24, 30, 36, 42, 48, 54, and 60 months. MDS-UPDRS, Movement Disorder Society Unified Parkinson’s Disease Rating Scale; MoCA, Montreal Cognitive Assessment; SCOPA-AUT, Scales for Outcomes in Parkinson’s Disease-Autonomic.
