## Supplementary Table 3 for "Parkinson’s Progression Markers Initiative: A Milestone-Based Strategy to Monitor PD Progression"

**Supplementary Table 3. Stability (overall and by domain) of milestone criteria at subsequent annual visits**

|  | **Domain first reached** | | | | | | |
| --- | --- | --- | --- | --- | --- | --- | --- |
| **Variable** | **Overall (any domain)** | **Cognition domain** | **Functional dependence** | **Autonomic dysfunction** | **Walking and balance** | **Motor complications** | **ADLs domain** |
| **Met criteria by 60-month visit**, n * | 166 | 53 | 45 | 41 | 25 | 19 | 17 |
| **Withdrew before next annual visit**, n | 14 | 3 | 6 | 1 | 4 | 2 | 3 |
| **N subsequent visits completed**, mean ** | 3.6 (1.8) | 3.5 (1.7) | 3.6 (1.8) | 3.6 (1.6) | 3.9 (2.1) | 3.1 (1.7) | 2.9 (2.0) |
| Median (min, max) ** | 3 (1, 8) | 3 (1, 7) | 3 (1, 7) | 3.5 (1, 7) | 4 (1, 7) | 3 (1, 6) | 2 (1, 8) |
| **Met *any* domain criteria at *any* visit**, n (%) ** | 125 (82%) | 43 (86%) | 33 (85%) | 31 (78%) | 21 (100%) | 12 (71%) | 12 (86%) |
| **Met *any* domain criteria at *next* visit**, n (%) ** | 84 (55%) | 27 (54%) | 24 (62%) | 21 (53%) | 18 (86%) | 10 (59%) | 9 (64%) |
| **Met *same* domain criteria at *next* visit**, n (%) ** | — | 25 (50%) | 17 (44%) | 15 (38%) | 12 (57%) | 5 (29%) | 6 (43%) |

Reflects data collected at *annual* visits only and includes all corresponding data captured in the PPMI database as of June 30, 2020 (maximum duration of follow-up: 96 months). *Indicates how many participants met the corresponding criteria at the *initial* event (i.e., first visit at which criteria for at least one milestone from any domain were met). **Calculations exclude anyone who withdrew before completing another follow-up. ADLs, activities of daily living.
