## Supplementary figures and images for "Parkinson’s Progression Markers Initiative: A Milestone-Based Strategy to Monitor PD Progression"

### Supplementary Figure 1

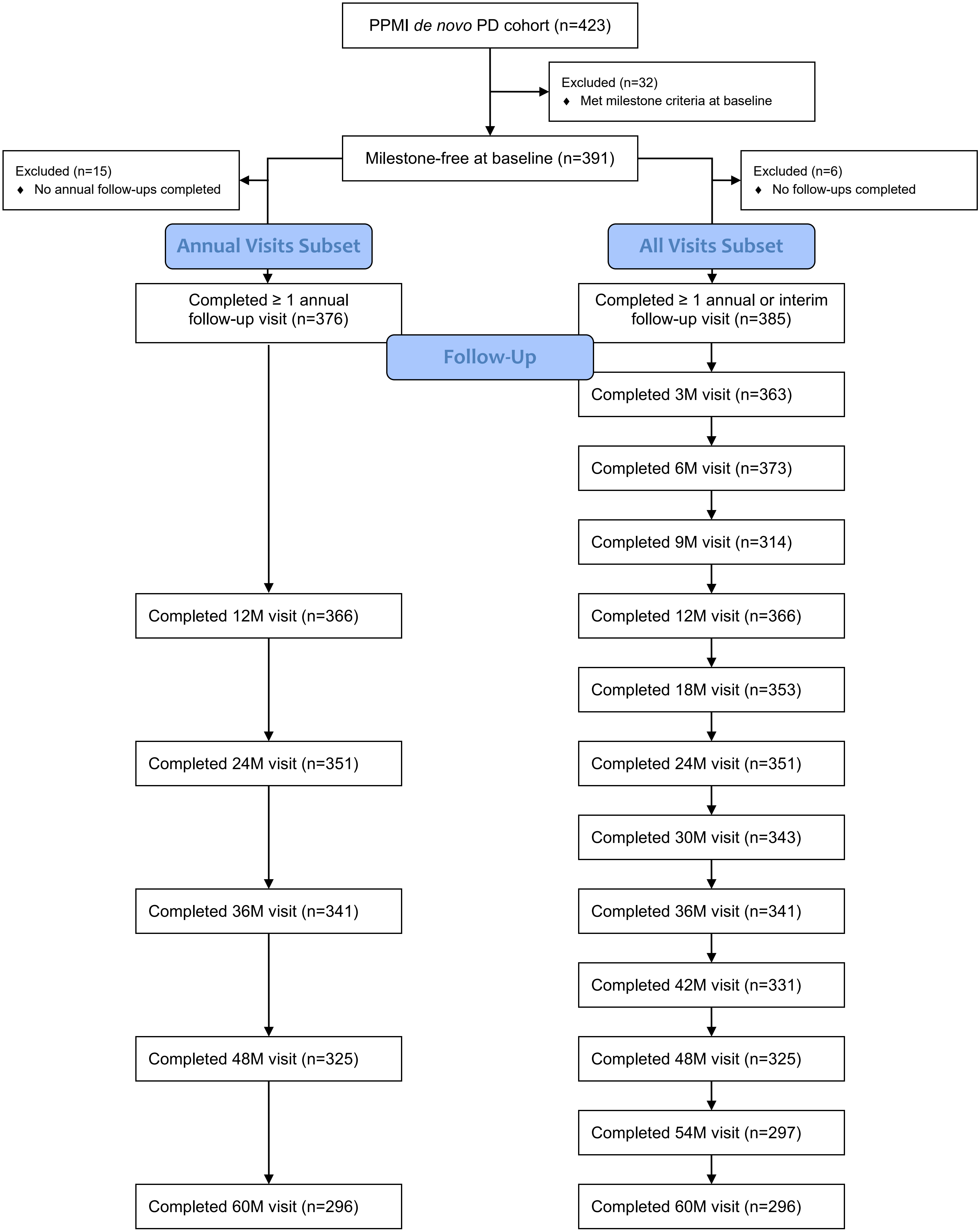
